# Commercial COVID-19 serial seroconversion panel for validation of SARS-CoV-2 antibody assays

**DOI:** 10.1101/2020.09.08.20190256

**Authors:** Francisco Belda, Robin Cherenzia, Michael Crowley

## Abstract

**Background:** Seroconversion panels (SCP) are an important tool for investigating antibody responses and developing serological assays. A SCP was generated from a single SARS-CoV-2 positive plasma donor over 87 days.

**Methods:** This SCP was tested against five SARS-CoV-2 antibody tests (IgG, IgM and total Ig). All test kits utilized recombinant antigens that are specific to SARS-CoV-2.

**Results:** The SCP showed IgG responses for SARS-CoV-2 after day 50. IgM levels peaked on day 50 (prior to IgG) and declined in subsequent samples.

**Conclusion:** This SCP is a useful tool for validation of SARS-CoV-2 antibody assays.

## Introduction

The emergence of the SARS-CoV-2 virus in Wuhan, China (December 2019) [1] and the resulting infection (COVID-19) has resulted in a global pandemic with significant morbidity and mortality [2]. The SARS-CoV-2 virus has been completely sequenced and shows substantial homology with SARS-CoV-1, the virus that causes severe acute respiratory syndrome (SARS) [3].

As yet, no specific treatment has been identified to prevent or treat COVID-19. Public health efforts are focused on determination of the prevalence and containment of the spread of COVID-19. Serologic testing is key to providing data not only for estimation of prevalence but also tracking and containment of the virus in the population. Serologic assays for SARS-CoV-2 antibodies can differ in sensitivity and specificity as well as window period of detection. One important tool in studying antibody response for window period detection and validation of an assay is a seroconversion panel collected over time (pre and post-infection).

Seroconversion panels have been used for multiple purposes including antibody assay development, process and product validation and quality control. Regulatory authorities may require or recommend validation with seroconversion panels. For example, the U.S. Food and Drug Administration (FDA) Guidance for Hepatitis A Virus Serological Assays recommends incorporation of seroconversion panels in the validation testing plan in order to assess the appearance of the analyte and the waning of IgM over time [4]. Available seroconversion panels provide patient samples over time for a number of viral infectious diseases including human immunodeficiency virus (HIV), hepatitis B virus (HBV), hepatitis C virus (HCV) and hepatitis A virus (HAV).

Currently, there is no commercial seroconversion panel available for SARS-CoV-2. A seroconversion panel was identified and generated from a single COVID-19 plasma donor. This serial panel represented pre-infection, infection and convalescence over 87 days. Characterization of the antibody response over time was evaluated by enzyme-linked immunosorbent assays (ELISA) and chemiluminescent assays (CLIA).

## Material and Methods

Five anti-SARS-CoV-2 antibody tests were used in this study to characterize a COVID-19 seroconversion panel (COVID-19 Seroconversion Panel: CVD19SCP, Access Biologicals, Vista, CA, USA). The antibody tests used were: the Gold Standard™ Diagnostics SARS-CoV-2 IgG ELISA and SARS-CoV-2 IgM ELISA test kits (Gold Standard™ Diagnostics (GSD), Davis, CA, USA; CE-IVD certified immunoassays, emergency use authorization (EUA) submission pending), Vitros® anti-SARS-CoV-2 IgG and anti-SARS-CoV-2 total Ig tests (Vitros® Immunodiagnostic Products, Ortho-Clinical Diagnostics, Inc., Rochester, NY, USA; EUA approved) and Liaison® SARS-CoV-2 S1/S2 IgG assay (Diasorin, Inc., Saluggia, Italy; EUA approved). All the antibody test kits were used according to the manufacturer’s instructions. All three test kits utilize recombinant antigens specific to SARS-CoV-2 (GSD, antigenic proteins for Ig G and Ig M; Vitros®, spike protein for Ig G and spike protein S1 for Ig M and Diasorin, spike protein S1 and S2 for Ig G [data obtained from the kits’ manufacturing instructions]). Although the exact sequences of the antigens are not specified, they were independently created and presumed to be non-identical.

The seroconversion panel consisted of 14 vials of 1.0 mL each of human plasma collected from a single COVID-19 donor. The samples were collected for 87 days (from 4^th^ of March to 29^th^ of May) over the course of the infection at an FDA-licensed plasma donor center (Saturn Biomedical, Indianapolis, IN, USA). The samples were collected with informed consent, under an approved IRB protocol (Advarra, Columbia MD, USA) and in compliance with all applicable regulatory guidelines. The preservative-free plasma samples were collected in 4% sodium citrate and aseptically filtered. Samples were stored at −20° C until use. Prior to use the samples were thawed at room temperature and gently mixed by inversion.

Plasma from this single donor was screened and found to be negative for syphilis and antibodies to HIV-1/2 HCV and non-reactive for hepatitis B surface antigen (HBsAg). In addition, non-reactive results were obtained for HIV-1/2 RNA, HBV DNA and HCV RNA using FDA-approved nucleic acid test assays. All donor plasma samples were also non-reactive for SARS-CoV-2 using a nucleic acid transcription-mediated amplification SARS-CoV-2 assay (Procleix® assay and Procleix Panther^®^ system, Grifols Diagnostic Solutions Inc., San Diego, CA). This SARS-CoV-2 assay is a qualitative in vitro nucleic acid transcription-mediated amplification test for the detection of the SARS-CoV-2 RNA in plasma, serum and respiratory specimens. Based on probit analysis, the 95% limits of detection of heat-inactivated SARS-CoV-2 (NR-52286, BEI Resources, Manassas, VA) diluted in K2EDTA plasma was estimated to be 10.7 copies/mL with 95% fiducial limits of 8.7 - 14.1 copies/mL [5].

## Results

As shown in Figure 1, the seroconversion panel showed positive IgG responses for SARS-CoV-2 at times ≥ day 50 with all three assay kits. Each assay kit expressed the results in arbitrary units. In order to allow comparison across the assay kits, results from all three kits were expressed on a 100-MEDRXIV/2020/190256 point scale (see Figure 1). For all three assays, based on the manufacturer’s limits of detection, all values prior to day 50 were considered non-reactive.

**Figure 1.**
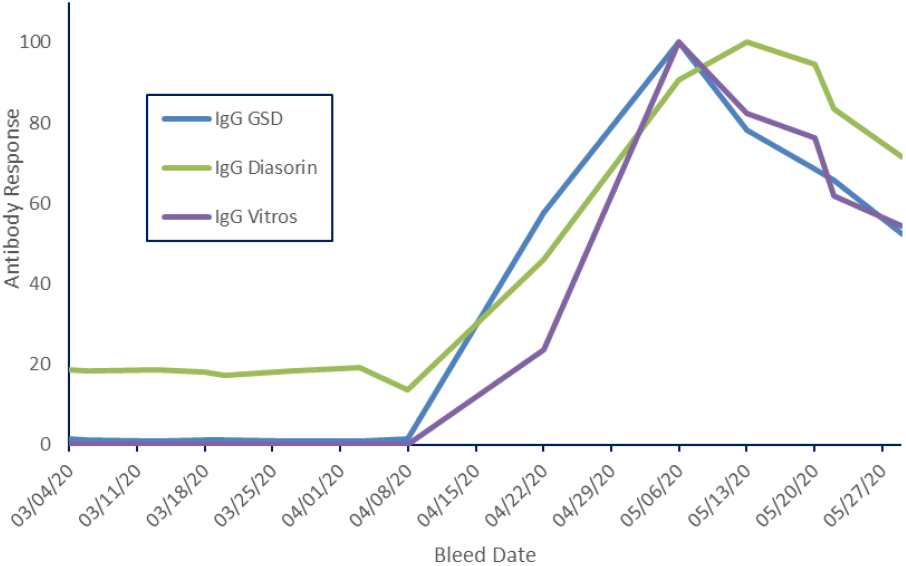
Levels of anti-SARS-CoV-2 IgG antibodies in a seroconversion panel as measured by three different assays. Each assay assigned arbitrary units to IgG levels so levels were normalized to a 100 point scale to allow comparison across assays.

IgG levels increased in all three assays at times ≥ day 50. Two of the assays detected peak IgG levels at day 64 and the other at day 71. After the peak response was seen, all three kits detected decreased anti-SARS-CoV-2 IgG levels over the remainder of the sample period. At 87 days, the anti-SARS-CoV-2 IgG levels were 60-70% of the peak level. The GSD assay kit measured anti-SARS-CoV-2 IgM as well as IgG.

Figure 2 shows the timeline for development of IgM and IgG for this assay. Antibody levels in this donor behave as expected; anti-SARS-CoV-2 IgM levels peaked prior to IgG levels (on day 50) and declined over the time course of the study. At 87 days IgM levels have fallen below the manufacturers cut-off and the sample would be considered non-reactive for IgM.

**Figure 2.**
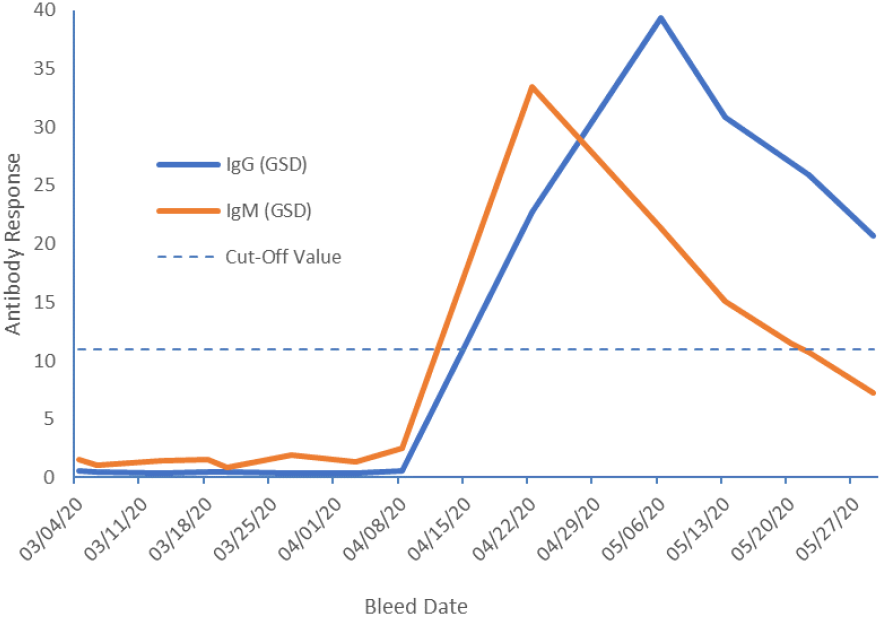
Anti-SARS-CoV-2 IgG and IgM antibodies measured in a SARS-CoV-2 seroconversion panel using Gold Standard^SM^ Diagnostics (GSD) kits. Values are expressed on an arbitrary scale specified by the manufacturer. Values above 11 are considered reactive (cut-off value). Values between 9 and 11 were considered equivocal and values below 9 were considered non-reactive.

The Vitros® assay measured total anti-SARS-CoV-2 Ig as well as IgG (Figure 3). Overall, total anti-SARS-CoV-2 Ig levels increased throughout the assay period peaking on day 87. Expressed relative to anti-SARS-CoV-2 IgG, total Ig was much higher than IgG.

**Figure 3.**
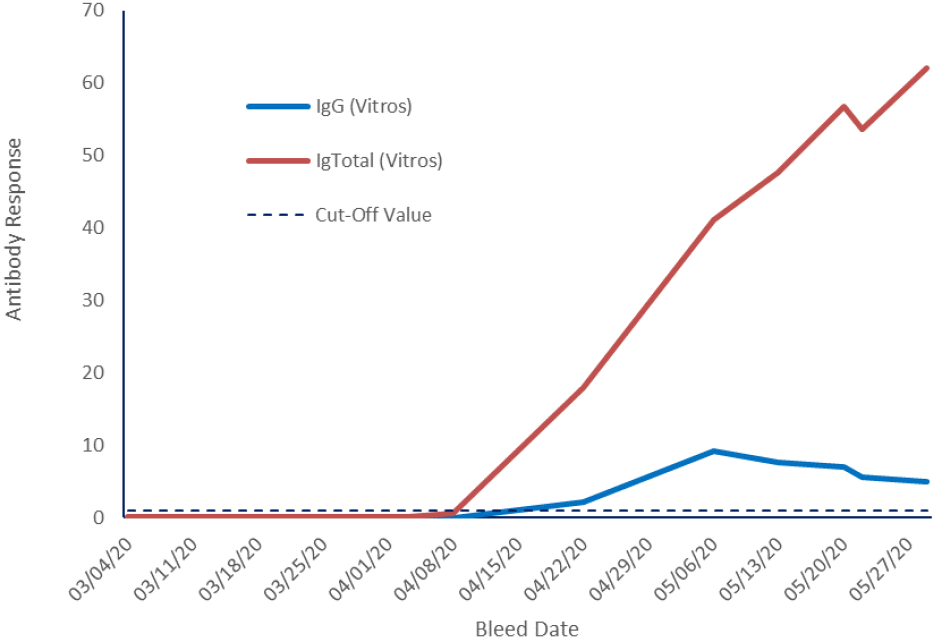
Anti-SARS-CoV-2 total immunoglobulins (Ig Total) and IgG measured in a SARS-CoV-2 seroconversion panel using Vitros® kits. Values were expressed on an arbitrary scale relative specified by the manufacturer. For this assay, values ≥ 1.0 were considered reactive (cut-off value) and values < 1.0 were considered non-reactive for SARS-CoV-2

## Discussion

In this study, a seroconversion panel obtained from a single patient was tested against three commercially available test kits for detecting anti-SARS-CoV-2 antibodies. This seroconversion panel is one of the first available for SARS-CoV-2. The advantage of this characterized seroconversion panel is the expansive time period (samples taken over almost 90 days) providing an extensive record of seroconversion in this single donor. This is a much longer time course than most seroconversion panels [6-8].

Similar results were obtained with all three assay kits which employ different recombinant SARS-CoV-2 antigens. Detection of anti-SARS-CoV-2 IgG antibodies showed the same time course in all three assays. This time course from infection or symptom development to antibody generation has been found to be highly variable between patients [9]. The MEDRXIV/2020/190256 time course for antibody generation in a different patient could be substantially different.

In this study, a decrease in IgG levels was observed after a peak at 64-71 days. IgG levels decreased to 60-70% of the peak level. Other studies have not shown this decrease in IgG at later time points, but these studies were shorter in duration which may account for the difference. In addition, IgM and IgG showed the expected temporal relationship in this seroconversion panel, i.e., IgM peaked prior to IgG and declined more steeply than IgG similar to the findings in other studies [10-12].

In conclusion, this seroconversion panel is a useful tool for developers of SARS-CoV-2 antibody detection assays, as well as for a proper validation of existing serological assays and as part of the decision making when choosing the most reliable serological kit for a specific application. SARS-CoV-2 antibody detection is now and will continue to be a valuable tool for detection of COVID-19 immune responses especially in asymptomatic and convalescent patients.

## Data Availability

Data referred to in the manuscript are available from the corresponding author upon reasonable request.

## Acknowledgements

Michael K James, PhD. (Grifols) is acknowledged for medical writing and Jordi Bozzo, PhD., CMPP (Grifols) is acknowledged for editorial support in the preparation of this manuscript. Contributions from Norbert Piel, Sansan Lin, Rodrigo Gajardo and Jerry A. Holmberg (Grifols) who provided their expert opinions are also acknowledged.

